# Prescribing of antibiotics and other drugs to children from birth to age 5 in the United States: an observational study

**DOI:** 10.1101/2021.11.17.21266458

**Authors:** Stephen M. Kissler, Bill Wang, Ateev Mehrotra, Michael Barnett, Yonatan H. Grad

## Abstract

**Objectives:** To inform efforts to reduce pediatric antibiotic use, we measured cumulative pediatric prescriptions for antibiotics and non-antibiotics and how this varies across geography and patient subgroups.

**Design:** Observational study.

**Setting:** United States, 2008-2018.

**Participants:** 207,814 children under age 5 born in the United States between 2008 and 2013 with private medical insurance coverage.

**Interventions:** None.

**Main outcome measures:** Study outcomes included (1) the cumulative number of prescriptions received per child by age 5, (2) the proportion of these prescriptions that were attributable to respiratory infections, (3) the proportion of children who received at least one prescription by age 5, and (4) the fraction of total prescriptions received by the top 20% of prescription recipients.

**Results:** Children received a mean of 8.21 (95% confidence interval [CI] (8.19, 8.22)) prescriptions for antibiotics and 9.81 (95% CI 9.80, 9.82) prescriptions for non-antibiotics by age five. Most antibiotic prescriptions (64%, 95% CI 63, 65) and many non-antibiotic prescriptions (25%, 95% CI 24, 26) were associated with outpatient visits for respiratory infections. By age 5, 93.8% (95% CI 93.4, 94.2) of children had received at least one antibiotic prescription while 88.3% (95% CI 87.9, 88.7) had received at least one prescription for a non-antibiotic. The top 20% of antibiotic prescription recipients accounted for 50.6% of all antibiotic prescriptions, and the top 20% of non antibiotic prescription recipients accounted for 64.2% of all non-antibiotic prescriptions. Relative to other regions, the South featured higher prescribing rates and earlier time to first prescription.

**Conclusions:** Children in the US receive a substantial number of antibiotics and other prescription drugs early in their lives, largely related to respiratory infections.

## Introduction

Antibiotics are critical therapeutics, yet their effectiveness is threatened by high levels of antibiotic consumption^1^ leading to antibiotic resistance. Children under age 5 receive antibiotics at a higher rate than any other age group in the United States^2^ and thus are an important target population for reducing antibiotic consumption.^2^ Beyond contributing to resistance, pediatric antibiotic consumption may affect the development of a healthy gut microbiome^3^ and is associated with childhood obesity.^4^ However, little is known about the individual-level patterns of pediatric antibiotic prescribing in the US. Cross-sectional studies have measured the mean antibiotic consumption rates by age group^5,6^ and determined that pediatric antibiotic prescriptions are frequently prompted by respiratory infections,^7,8^ yet such studies do not capture the cumulative exposure of individual children to antibiotics during the first years of life, nor do they quantify the degree of variation in antibiotic consumption among children, which may be a key variable determining the evolution and proliferation of antibiotic resistance.^9^

Since antibiotic prescriptions are frequently prompted by respiratory infections, respiratory disease prevention (*e*.*g*., through vaccination) offers a promising approach for reducing antibiotic consumption.^10,11^ Quantifying the potential benefits of such interventions requires a detailed description of pediatric antibiotic consumption patterns. Additionally, respiratory disease prevention could reduce consumption of other prescription drugs used to manage the symptoms and sequalae of respiratory infections. However, little is known about the rate at which children receive non-antibiotic prescriptions and the share of these non-antibiotic prescriptions that are also associated with respiratory disease.

To address these evidence gaps in early life prescription drug use, we estimated the cumulative number of prescriptions for antibiotics and non-antibiotics received before age 5 for children in the US with private medical insurance and the fraction of these prescriptions that were attributable to respiratory infections.

## Methods

### Study sample and data sources

Prescribing data were obtained from pharmacy claims records aggregated in the Truven MarketScan database.^12^ These data represent a convenience sample of 19.1–24.3 million individuals (5.9–7.6% of the US population) with private medical insurance coverage, which varies based on insurance enrollment by month. Each pharmacy claim includes the fill date, the National Drug Code (NDC) identifier, a drug class (**Supplementary Table 1**), and the patient’s age, sex, state, and metropolitan statistical area (MSA)^13^ of residence. The database also includes outpatient and inpatient claims with a primary diagnosis coded using the International Classification of Diseases, revision 9 (ICD9) or 10 (ICD10). We restricted the sample to children continuously enrolled through age 5 who were born between 1 January 2008 and 31 December 2013, following previously described methods^14^ (*n* = 207,814) (**Table 1, Supplementary Table 2**).

**Table 1.**
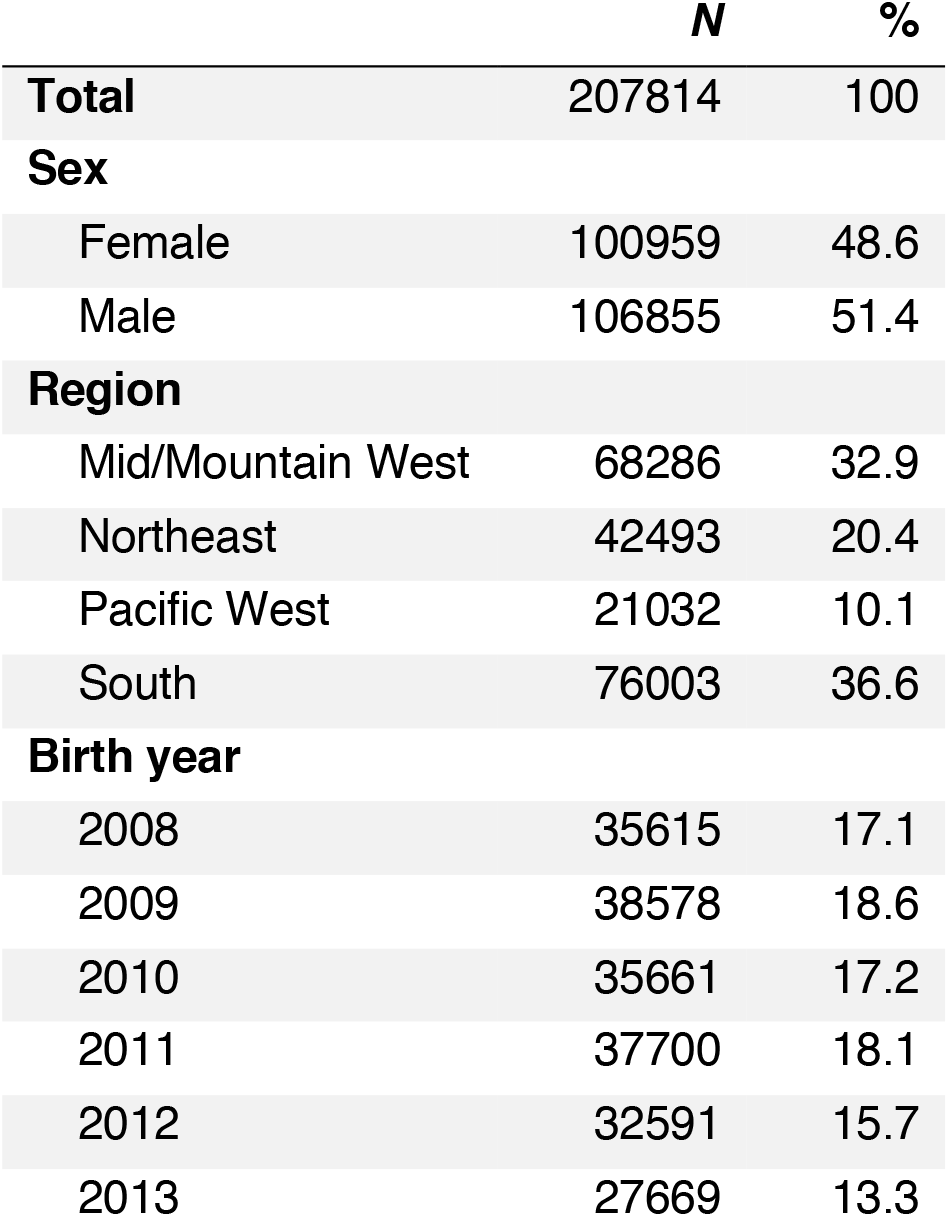
Summary of the study population. Number of individuals (*N*) and percent of the total cohort (%) by sex, region, and birth year.

|  | *N* | % |
| --- | --- | --- |
| **Total** | 207814 | 100 |
| **Sex** |  |  |
| Female | 100959 | 48.6 |
| Male | 106855 | 51.4 |
| **Region** |  |  |
| Mid/Mountain West | 68286 | 32.9 |
| Northeast | 42493 | 20.4 |
| Pacific West | 21032 | 10.1 |
| South | 76003 | 36.6 |
| **Birth year** |  |  |
| 2008 | 35615 | 17.1 |
| 2009 | 38578 | 18.6 |
| 2010 | 35661 | 17.2 |
| 2011 | 37700 | 18.1 |
| 2012 | 32591 | 15.7 |
| 2013 | 27669 | 13.3 |

### Identifying outpatient visits, linking to prescriptions and diagnoses

Prescriptions were linked to the most recent outpatient visit within 7 days. If a prescription was linked with an outpatient claim that listed a respiratory infection in either the first or second diagnosis field, then the prescription was deemed to be due to a respiratory infection. Respiratory infection diagnoses were identified using the Agency for Health Research and Quality Clinical Classification Software (CCS).^15^ (list in **Supplementary Table 3)**.

### Study outcomes and covariates

We examined four main outcomes: (1) the mean cumulative number of prescriptions received per child by age 5, (2) the proportion of these prescriptions that were attributable to respiratory infections, (3) the proportion of children who had received at least one prescription by age 5, and (4) the fraction of total prescriptions that were received by the top 20% of prescription recipients.

### Statistical analysis

To extrapolate to a population more representative of children in the United States under age 5, we weighted each claim according to the child’s sex and state. Weights were defined by the ratio of (a) the number of children under 5 in that state/sex group in the US population, according to the 2010 US census,^16^ and (b) the number of children in the MarketScan data in the same state/sex group (**Supplementary Table 2**). To estimate the number of prescriptions in the US population under age 5, we multiplied the visit and prescription counts for each person in the MarketScan data by these weights.

To assess geographic variation, we examined prescribing by census region (Northeast, South, Mid/Mountain West, and Pacific West; **Supplementary Table 4**) and by metropolitan statistical area (MSA),^13^ a designation assigned by the US Census Bureau that roughly corresponds to a city. For the MSA-level analysis, we excluded MSAs that had fewer than 100 cohort members. These remaining 219 MSAs included 85% (*n =* 177,677) of the study population.

Analyses were conducted using R, version 3.6.2. Confidence intervals were calculated as exact 95% confidence intervals for Poisson means. This study was determined as not human subjects research by the Harvard T.H. Chan School of Public Health Institutional Review Board (Protocol #IRB20-0130).

## Results

Among 207,814 individuals born in 2008–2013 in the study sample, children received a mean of 8.21 (95% CI 8.19, 8.22) prescriptions for antibiotics and 9.81 (95% CI 9.80, 9.82) prescriptions for non-antibiotics by age five (**Figure 1A, Supplementary Table 4**). Antibiotic prescribing rates were highest between 6 months and 2 years of age, when the mean prescribing rate exceeded 0.15 antibiotic prescriptions per child per 30 days (**Figure** 1**A**, **Supplementary Figure 1**). Most antibiotic prescriptions (64%, 95% CI 63, 65) and a substantial minority of non-antibiotic prescriptions (25%, 95% CI 24, 26) were associated with outpatient visits for respiratory infections.

**Figure 1.**
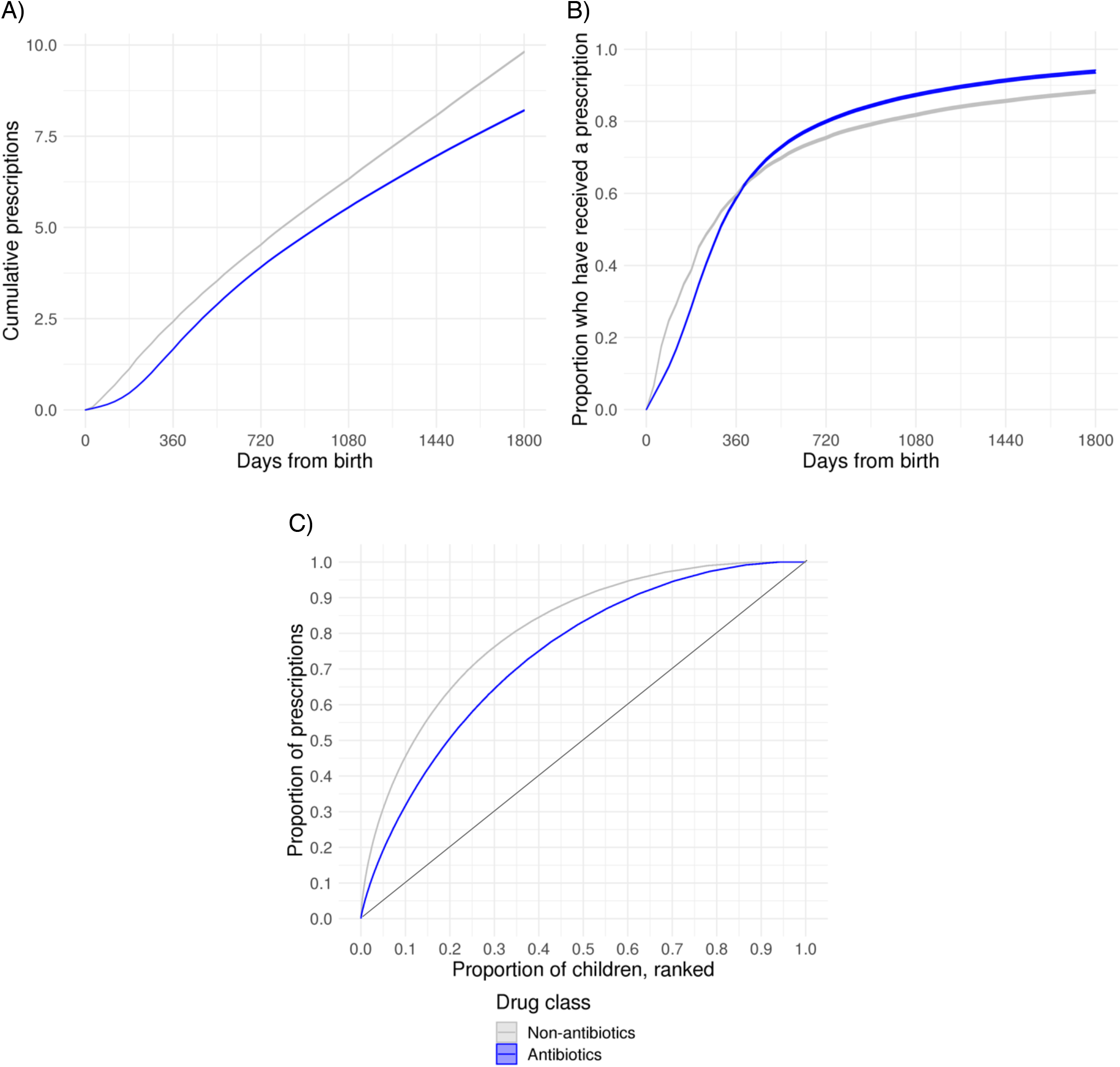
Patterns in prescribing of antibiotics and non-antibiotics to children under age 5 in the United States. (A) Mean cumulative number of prescriptions received per child between birth and age 5. (B) Proportion of children who have received at least one prescription between birth and age 5. (C) Lorenz curves depicting the proportion of prescriptions (vertical axis) attributable to different proportions of children (horizontal axis), ranked from the most frequent users (left) to the least frequent (right). The thin grey line depicts the total equality scenario, where k% of children are responsible for k% of prescriptions. In all figures, lines depict means and bands depict 95% confidence intervals according to a two-tailed *t*-test for the mean.

Similar proportions of children received at least one antibiotic prescription (58.6%, 95% CI 58.3, 59.0) or at least one non-antibiotic prescription (59.5%, 95% CI 59.2, 59.8) by age 1 (**Figure** 1**B**, **Supplementary Table 6**). By age 5, 93.8% (95% CI 93.4, 94.2) of children had received at least one antibiotic prescription while 88.3% (95% CI 87.9, 88.7) had received at least one prescription for a non-antibiotic (**Figure** 1**B, Supplementary Table 6**).

Children varied widely in the number of prescriptions received (**Figure** 1**C**, **Supplementary Figure 2**). The top 20% of antibiotic recipients (*i*.*e*., those with the most prescriptions received by age 5) accounted for 50.6% of all antibiotic prescriptions. This variation was more pronounced for non-antibiotics: the top 20% of recipients accounted for 64.2% all non-antibiotic prescriptions (Figure 1**C**).

Cumulative prescriptions by age 5 for all drug classes were higher in MSAs in the South (antibiotics: 9.79 (95% CI 9.73, 9.83); non-antibiotics: 11.81 (95% CI 11.73, 11.88)) than in the rest of the country, such as the Pacific West (antibiotics: 6.07 (95% CI 5.87, 6.17); non-antibiotics: 7.76 (95% CI 7.62, 7.87)) (**Figure 2A, Supplementary Table 7**). The penetrance of prescribing was also higher in the South. For example, the proportion of children who received at least one antibiotic prescription by age 5 was 0.96 (95% CI 0.95, 0.97) in the South *vs*. 0.91 (95% CI 0.90, 0.92) in the Pacific West (**Figure 2B, Supplementary Table 7**). The top 20% of prescription recipients accounted for a smaller share of prescriptions in the South than in in the rest of the country: these children accounted for 46% (95% CI 45, 47) of antibiotic prescriptions in the South *vs*. 50% (95% CI 49, 51) in the Pacific West (**Figure 2C, Supplementary Table 7**). More non-antibiotic prescriptions were associated with respiratory infections in the South relative to the rest of the US (proportion of non-antibiotic prescriptions associated with respiratory infections in the South: 0.30 (95% CI 0.29, 0.31); in the Pacific West: 0.22 (95% CI 0.21, 0.23)), while the proportion of antibiotic prescriptions associated with respiratory infections were similar across regions (**Figure 2D, Supplementary Table 7**).

**Figure 2.**
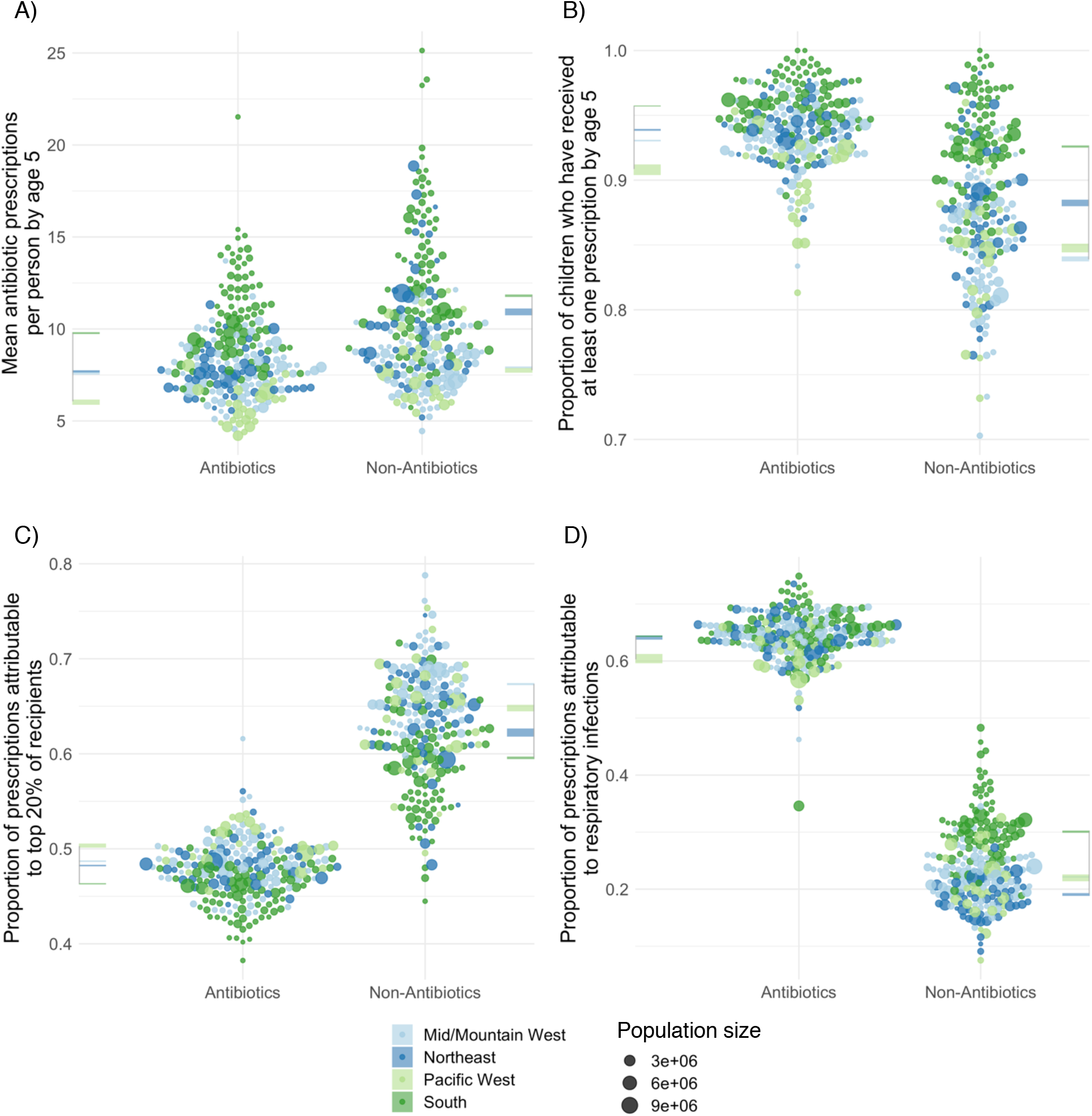
Geographic distribution of prescribing for children under age 5 in the United States by Metropolitan Statistical Area and drug class. Bee swarm plots depicting the distribution of (A) the mean cumulative number of prescriptions received per child between birth and age 5, (B) the proportion of children who have received at least one prescription between birth and age 5, (C) the proportion of prescriptions attributable to the top 20% of recipients, and (D) the proportion of prescriptions attributable to respiratory infections for the 219 largest Metropolitan Statistical Areas (MSAs) in the US, separated by antibiotics and non-antibiotics. Points represent Metropolitan Statistical Areas (MSAs), point size is proportional to the MSA’s population size, and colors represent the geographic region where the MSA is located. Colored horizontal bars depict the 95% confidence intervals for the MSA-level mean values as computed from 10,000 bootstrap samples. Regional means and confidence intervals are listed in **Supplementary Table 7**.

## Discussion

Children in the United States receive on average more than 8 antibiotic prescriptions in early childhood. This varies substantially across geography, with children in some MSAs receiving more than 15 antibiotic prescriptions by age 5. Prescribing patterns were similar for antibiotics and nonantibiotics, with high penetrance (most children receive at least one prescription by age 5) and substantial variation (fewer than 20% of children account for over 50% of prescriptions in both classes). Respiratory infections appear to be the major driver of antibiotic prescriptions and play an important role for non-antibiotic prescriptions.

While much of the focus on reducing antibiotic prescriptions has focused on stewardship, our results highlight how interventions to deter respiratory infections could be a promising strategy to reduce overall prescribing of antibiotics. The recent success of mRNA vaccines in the COVID-19 pandemic underscores the possibility of developing highly effective vaccines for other respiratory pathogens that could have the added benefit of reducing downstream antibiotic prescribing.

When designing strategies to address antibiotic resistance, inequalities in antibiotic prescribing rates across children may play a critical role. While large reductions in prescribing might be achieved by focusing on children who receive the most antibiotic prescriptions, children who receive relatively few antibiotics may contribute a greater share to overall resistance.^9^ Toward this end, efforts to document trends in antibiotic prescribing should measure changes in the distribution of prescriptions received across the population whenever possible, rather than just the mean.

Our results are broadly consistent with prior studies that have measured cumulative antibiotic prescribing in children in other high-income countries and, within the US, in more geographically restricted cohorts and in cross-sectional studies. In New Zealand, children received a mean of 8 antibiotic prescriptions before age 5 and 97% of children received at least one antibiotic by age 5.^17^ In a Philadelphia cohort, 69% of children received at least one antibiotic before age 2 for a mean of 2.3 antibiotic courses per child, in line with our findings.^4^ Prior cross-sectional studies have found higher antibiotic prescribing rates in the southeastern US relative to the rest of the country.^5,19^ However, our national estimates of antibiotic prescribing rates in children under 5 are higher than what can be extrapolated from these studies. According to these studies, annual antibiotic prescribing rates in US children under 5 lie between 562 and 1,287 annual prescriptions per 1,000;^5,6^ a simple extrapolation of these rates to 5 years of age would yield 2.8–6.4 prescriptions per child by age 5. Our estimates of cumulative antibiotic prescribing are also much lower than what has been observed in some low- and middle-income countries, where children receive an estimated 24.5 antibiotic prescriptions before age 5.^18^

Limitations to this study include possible bias introduced by convenience sampling and the exclusion of individuals without insurance. The study sample was also likely to exclude the dependents of individuals who frequently change employers (and thus insurance providers), since these were less likely to be represented continuously for five years in the data. Another limitation is that we are unable to directly observe the indication for prescribing. We infer the indication by looking at associated diagnosis codes with recent visits proximal to the prescription, but this may misclassify some prescriptions. Additionally, we cannot distinguish between appropriate and inappropriate prescribing of antibiotics or non-antibiotics.

In conclusion, children in the US are exposed to antibiotics and other prescription drugs frequently and at an early age. Reducing antibiotic prescribing rates remains a public health priority. If these reductions in prescribing are achieved by improving disease prevention through measures such as vaccines, we may attain the additional benefit of reducing prescriptions of other drugs, further reducing healthcare costs and avoiding potential side effects from excess drug exposure.

## Data Availability

All data produced in the present study are available upon reasonable request to the authors

---

**Supplementary Table 1.** Most common prescription classes in the study cohort. Columns list the number of filled prescriptions (*N*) and percent of all prescriptions (%) by therapeutic class. Antibiotics are marked in bold. Only the therapeutic classes that comprise at least 1% of all prescriptions are listed, along with the remaining five antibiotic classes that make up less than 1% of all prescriptions.

| Therapeutic class | *N* | % |
| --- | --- | --- |
| **Antibiot, Penicillins** | 812342 | 20.7 |
| **Antibiot, Cephalosporin & Rel.** | 372469 | 9.5 |
| Adrenals & Comb, NEC | 338434 | 8.6 |
| Sympathomimetic Agents, NEC | 297946 | 7.6 |
| **Antiinfect, Antibiotics EENT** | 254843 | 6.5 |
| **Antibiot, Erythromycin&Macrolid** | 250593 | 6.4 |
| Antiinflam S/MM Agnts&Comb NEC | 155820 | 4 |
| Histamine (H2) Antagonist, NEC | 135287 | 3.5 |
| Antiinf S/MM,Antifungal & Comb | 133483 | 3.4 |
| Leukotriene Modifiers | 112060 | 2.9 |
| Antihistamines & Comb, NEC | 99784 | 2.5 |
| Multivit Prep,Multivit Fluorid | 85298 | 2.2 |
| Gastrointestinal Drug Misc,NEC | 79553 | 2 |
| **Antiinf S/MM,Antibiotic & Comb** | 65712 | 1.7 |
| Sulfonamides & Comb, NEC | 65404 | 1.7 |
| Fluoride Preparations, NEC | 57303 | 1.5 |
| Antiinflam Agents EENT, NEC | 55061 | 1.4 |
| Antivirals, NEC | 54031 | 1.4 |
| Antiemetics, NEC | 44705 | 1.1 |
| Anesthetics, Local EENT, NEC | 39294 | 1 |
| ... | ... | ... |
| **Antibiot, Antifungals** | 20812 | 0.5 |
| **Antibiotics, Misc** | 10978 | 0.3 |
| **Antibiot, Aminoglycosides** | 380 | 0 |
| **Antibiot, Tetracyclines** | 169 | 0 |
| **Antibiot, B-Lactam Antibiotics** | 12 | 0 |

**Supplementary Table 2.** Characteristics of the study population. Number of individuals (*N*), reference population sizes and population fractions from the 2010 US census, and weights applied to each claim, equal to the 2010 US Census population fraction divided by the number of individuals *N*.

| State | Sex | *N* | Population size,<br>2010 US Census | Population fraction,<br>2010 US Census | Weight (x 10<sup>-6</sup>) |
| --- | --- | --- | --- | --- | --- |
| Alabama | F | 526 | 148,372 | 0.0074 | 14.07 |
| Alabama | M | 657 | 154,287 | 0.0077 | 11.72 |
| Alaska | F | 117 | 24,646 | 0.0012 | 10.51 |
| Alaska | M | 120 | 26,714 | 0.0013 | 11.11 |
| Arizona | F | 1970 | 226,542 | 0.0113 | 5.74 |
| Arizona | M | 2055 | 236,518 | 0.0118 | 5.74 |
| Arkansas | F | 438 | 95,376 | 0.0048 | 10.86 |
| Arkansas | M | 456 | 100,110 | 0.005 | 10.95 |
| California | F | 3212 | 1,244,216 | 0.0621 | 19.32 |
| California | M | 3075 | 1,300,849 | 0.0649 | 21.1 |
| Colorado | F | 1437 | 166,096 | 0.0083 | 5.77 |
| Colorado | M | 1547 | 173,902 | 0.0087 | 5.61 |
| Connecticut | F | 1339 | 100,161 | 0.005 | 3.73 |
| Connecticut | M | 1352 | 105,318 | 0.0053 | 3.89 |
| Delaware | F | 1478 | 27,534 | 0.0014 | 0.93 |
| Delaware | M | 1587 | 28,476 | 0.0014 | 0.9 |
| Florida | F | 4793 | 528,479 | 0.0264 | 5.5 |
| Florida | M | 5043 | 552,027 | 0.0275 | 5.46 |
| Georgia | F | 7255 | 336,710 | 0.0168 | 2.32 |
| Georgia | M | 7719 | 351,811 | 0.0176 | 2.27 |
| Idaho | F | 348 | 58,198 | 0.0029 | 8.34 |
| Idaho | M | 374 | 60,968 | 0.003 | 8.13 |
| Illinois | F | 4827 | 413,907 | 0.0206 | 4.28 |
| Illinois | M | 5019 | 431,211 | 0.0215 | 4.29 |
| Indiana | F | 2054 | 212,592 | 0.0106 | 5.16 |
| Indiana | M | 2200 | 221,612 | 0.0111 | 5.03 |
| Iowa | F | 1146 | 96,793 | 0.0048 | 4.21 |
| Iowa | M | 1195 | 101,153 | 0.005 | 4.22 |
| Kansas | F | 988 | 98,435 | 0.0049 | 4.97 |
| Kansas | M | 1063 | 100,765 | 0.005 | 4.73 |
| Kentucky | F | 1312 | 136,071 | 0.0068 | 5.17 |
| Kentucky | M | 1320 | 142,230 | 0.0071 | 5.38 |
| Louisiana | F | 795 | 149,242 | 0.0074 | 9.37 |
| Louisiana | M | 881 | 155,698 | 0.0078 | 8.82 |
| Maine | F | 258 | 33,622 | 0.0017 | 6.5 |
| Maine | M | 241 | 36,243 | 0.0018 | 7.5 |
| Maryland | F | 848 | 178,868 | 0.0089 | 10.52 |
| Maryland | M | 875 | 186,225 | 0.0093 | 10.62 |
| Massachusetts | F | 3849 | 180,330 | 0.009 | 2.34 |
| Massachusetts | M | 4033 | 187,996 | 0.0094 | 2.33 |
| Michigan | F | 6975 | 300,463 | 0.015 | 2.15 |
| Michigan | M | 7151 | 314,019 | 0.0157 | 2.19 |
| Minnesota | F | 1642 | 173,098 | 0.0086 | 5.26 |
| Minnesota | M | 1814 | 179,512 | 0.009 | 4.94 |
| Mississippi | F | 1607 | 101,976 | 0.0051 | 3.17 |
| Mississippi | M | 1641 | 106,515 | 0.0053 | 3.24 |
| Missouri | F | 3163 | 188,870 | 0.0094 | 2.98 |
| Missouri | M | 3513 | 197,907 | 0.0099 | 2.81 |
| Montana | F | 140 | 29,191 | 0.0015 | 10.4 |
| Montana | M | 135 | 30,531 | 0.0015 | 11.28 |
| Nebraska | F | 571 | 63,576 | 0.0032 | 5.55 |
| Nebraska | M | 587 | 65,834 | 0.0033 | 5.6 |
| Nevada | F | 654 | 91,863 | 0.0046 | 7.01 |
| Nevada | M | 735 | 96,732 | 0.0048 | 6.57 |
| New Hampshire | F | 626 | 35,525 | 0.0018 | 2.83 |
| New Hampshire | M | 681 | 36,772 | 0.0018 | 2.69 |
| New Jersey | F | 3365 | 267,716 | 0.0134 | 3.97 |
| New Jersey | M | 3667 | 279,434 | 0.0139 | 3.8 |
| New Mexico | F | 620 | 70,353 | 0.0035 | 5.66 |
| New Mexico | M | 705 | 72,947 | 0.0036 | 5.16 |
| New York | F | 4367 | 566,415 | 0.0283 | 6.47 |
| New York | M | 4685 | 592,250 | 0.0295 | 6.31 |
| North Carolina | F | 3506 | 303,237 | 0.0151 | 4.31 |
| North Carolina | M | 3730 | 318,634 | 0.0159 | 4.26 |
| North Dakota | F | 77 | 20,473 | 0.001 | 13.26 |
| North Dakota | M | 92 | 21,319 | 0.0011 | 11.56 |
| Ohio | F | 6711 | 355,298 | 0.0177 | 2.64 |
| Ohio | M | 7346 | 369,367 | 0.0184 | 2.51 |
| Oklahoma | F | 757 | 125,707 | 0.0063 | 8.28 |
| Oklahoma | M | 743 | 130,917 | 0.0065 | 8.79 |
| Oregon | F | 2240 | 114,208 | 0.0057 | 2.54 |
| Oregon | M | 2447 | 119,993 | 0.006 | 2.45 |
| Pennsylvania | F | 2253 | 355,520 | 0.0177 | 7.87 |
| Pennsylvania | M | 2353 | 371,755 | 0.0185 | 7.88 |
| Rhode Island | F | 271 | 28,772 | 0.0014 | 5.3 |
| Rhode Island | M | 297 | 30,148 | 0.0015 | 5.06 |
| South Carolina | F | -- | 145,046 | 0.0072 | -- |
| South Carolina | M | -- | 150,860 | 0.0075 | -- |
| South Dakota | F | 217 | 27,395 | 0.0014 | 6.3 |
| South Dakota | M | 242 | 29,099 | 0.0015 | 6 |
| Tennessee | F | 2920 | 197,994 | 0.0099 | 3.38 |
| Tennessee | M | 3097 | 205,473 | 0.0103 | 3.31 |
| Texas | F | 9903 | 931,442 | 0.0465 | 4.69 |
| Texas | M | 10529 | 972,112 | 0.0485 | 4.61 |
| Utah | F | 1172 | 124,041 | 0.0062 | 5.28 |
| Utah | M | 1203 | 130,971 | 0.0065 | 5.43 |
| Vermont | F | 50 | 15,557 | 8.00E-04 | 15.52 |
| Vermont | M | 85 | 16,698 | 8.00E-04 | 9.8 |
| Virginia | F | 1701 | 247,543 | 0.0123 | 7.26 |
| Virginia | M | 1815 | 258,706 | 0.0129 | 7.11 |
| Washington | F | 1762 | 208,047 | 0.0104 | 5.89 |
| Washington | M | 1923 | 218,403 | 0.0109 | 5.67 |
| Washington DC | F | 36 | 15,959 | 8.00E-04 | 22.12 |
| Washington DC | M | 50 | 16,463 | 8.00E-04 | 16.43 |
| West Virginia | F | 167 | 50,532 | 0.0025 | 15.1 |
| West Virginia | M | 164 | 53,226 | 0.0027 | 16.19 |
| Wisconsin | F | 1924 | 173,266 | 0.0086 | 4.49 |
| Wisconsin | M | 1908 | 181,588 | 0.0091 | 4.75 |
| Wyoming | F | 106 | 18,153 | 9.00E-04 | 8.54 |
| Wyoming | M | 121 | 19,115 | 0.001 | 7.88 |

**Supplementary Table 3.** Clinical classification software (CCS) codes for respiratory infections. Columns list the CCS code for the ICD9 (2015) and ICD10 (2021) revisions.

| Condition | CCS9 | CCS10 |
| --- | --- | --- |
| Sinusitis | — | RSP001 |
| Pneumonia | 122 | RSP002 |
| Influenza | 123 | RSP003 |
| Tonsillitis | 124 | RSP004 |
| Bronchitis (acute) | 125 | RSP005 |
| Upper respiratory infection (other) | 126 (Includes Sinusitis) | RSP006 |
| Otitis media | 92 | EAR001 |

**Supplementary Table 4.** Regions with constituent HHS categories and states. HHS categories^20^ consist of groupings of states that are further grouped into the four regions in our analysis.

| Region | HHS categories | States |
| --- | --- | --- |
| Northeast | 1, 2, 3 | Connecticut, Delaware, Maine, Maryland, Massachusetts, New Hampshire, New Jersey, New York, Pennsylvania, Rhode Island, Vermont, Virginia, Washington DC, West Virginia |
| South | 4, 6 | Alabama, Arkansas, Florida, Georgia, Kentucky, Louisiana, Mississippi, New Mexico, North Carolina, Oklahoma, South Carolina, Tennessee, Texas |
| Mid/Mountain West | 5, 7, 8 | Colorado, Illinois, Indiana, Iowa, Kansas, Michigan, Minnesota, Missouri, Montana, Nebraska, North Dakota, Ohio, South Dakota, Utah, Wisconsin, Wyoming |
| Pacific West | 9, 10 | Alaska, Arizona, California, Hawaii, Idaho, Nevada, Oregon, Washington |

**Supplementary Table 5.** Mean cumulative number of prescriptions received at various ages between birth and age 5 for antibiotics and non-antibiotics.

| Drug class | Age | Mean cumulative number of prescriptions |  |
| --- | --- | --- | --- |
| Non-Antibiotics | 1 year | 2.42 | (2.41, 2.43) |
|  | 2 years | 4.53 | (4.52, 4.53) |
|  | 5 years | 9.81 | (9.8, 9.82) |
| Antibiotics | 1 year | 1.66 | (1.66, 1.67) |
|  | 2 years | 3.91 | (3.9, 3.92) |
|  | 5 years | 8.21 | (8.19, 8.22) |

**Supplementary Table 6.** Percent of children who have received at least one prescription by age 5 for antibiotics and non-antibiotics.

| Drug class | Age | Percent of children who have received at least one prescription |  |
| --- | --- | --- | --- |
| Non-Antibiotics | 1 year | 59.5 | (59.2, 59.8) |
|  | 2 years | 75.4 | (75, 75.8) |
|  | 5 years | 88.3 | (87.9, 88.7) |
| Antibiotics | 1 year | 58.6 | (58.3, 59) |
|  | 2 years | 79.9 | (79.5, 80.3) |
|  | 5 years | 93.8 | (93.4, 94.2) |

**Supplementary Table 7.**
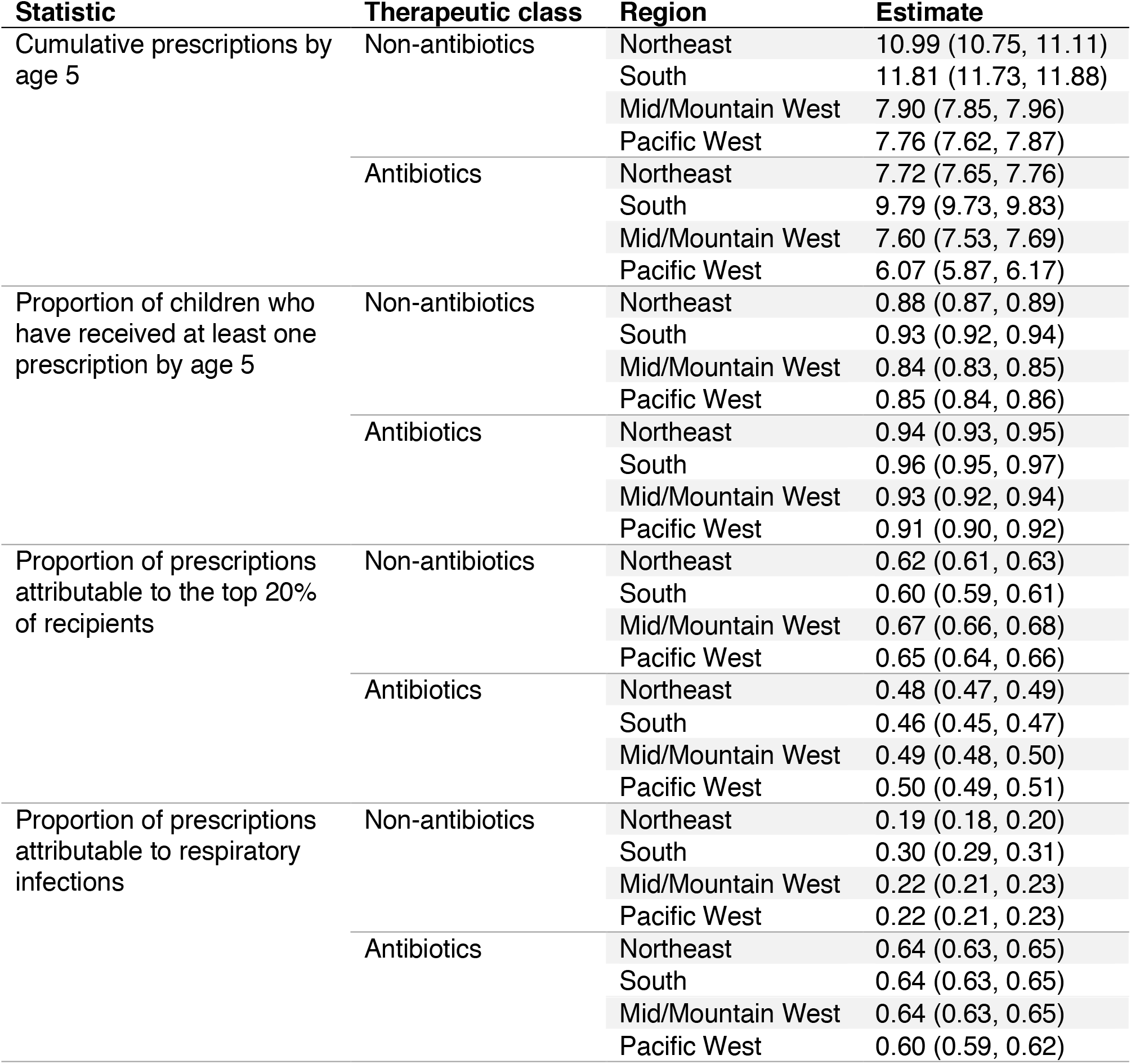
Regional estimates of key statistics by therapeutic class. Estimates represent regional means. Values in parentheses represent 95% confidence intervals as computed from 10,000 bootstrap samples.

| Statistic | Therapeutic class | Region | Estimate |
| --- | --- | --- | --- |
| Cumulative prescriptions by age 5 | Non-antibiotics | Northeast | 10.99 (10.75, 11.11) |
|  |  | South | 11.81 (11.73, 11.88) |
|  |  | Mid/Mountain West | 7.90 (7.85, 7.96) |
|  |  | Pacific West | 7.76 (7.62, 7.87) |
|  | Antibiotics | Northeast | 7.72 (7.65, 7.76) |
|  |  | South | 9.79 (9.73, 9.83) |
|  |  | Mid/Mountain West | 7.60 (7.53, 7.69) |
|  |  | Pacific West | 6.07 (5.87, 6.17) |
| Proportion of children who have received at least one prescription by age 5 | Non-antibiotics | Northeast | 0.88 (0.87, 0.89) |
|  |  | South | 0.93 (0.92, 0.94) |
|  |  | Mid/Mountain West | 0.84 (0.83, 0.85) |
|  |  | Pacific West | 0.85 (0.84, 0.86) |
|  | Antibiotics | Northeast | 0.94 (0.93, 0.95) |
|  |  | South | 0.96 (0.95, 0.97) |
|  |  | Mid/Mountain West | 0.93 (0.92, 0.94) |
|  |  | Pacific West | 0.91 (0.90, 0.92) |
| Proportion of prescriptions attributable to the top 20% of recipients | Non-antibiotics | Northeast | 0.62 (0.61, 0.63) |
|  |  | South | 0.60 (0.59, 0.61) |
|  |  | Mid/Mountain West | 0.67 (0.66, 0.68) |
|  |  | Pacific West | 0.65 (0.64, 0.66) |
|  | Antibiotics | Northeast | 0.48 (0.47, 0.49) |
|  |  | South | 0.46 (0.45, 0.47) |
|  |  | Mid/Mountain West | 0.49 (0.48, 0.50) |
|  |  | Pacific West | 0.50 (0.49, 0.51) |
| Proportion of prescriptions attributable to respiratory infections | Non-antibiotics | Northeast | 0.19 (0.18, 0.20) |
|  |  | South | 0.30 (0.29, 0.31) |
|  |  | Mid/Mountain West | 0.22 (0.21, 0.23) |
|  |  | Pacific West | 0.22 (0.21, 0.23) |
|  | Antibiotics | Northeast | 0.64 (0.63, 0.65) |
|  |  | South | 0.64 (0.63, 0.65) |
|  |  | Mid/Mountain West | 0.64 (0.63, 0.65) |
|  |  | Pacific West | 0.60 (0.59, 0.62) |

**Supplementary Figure 1.**
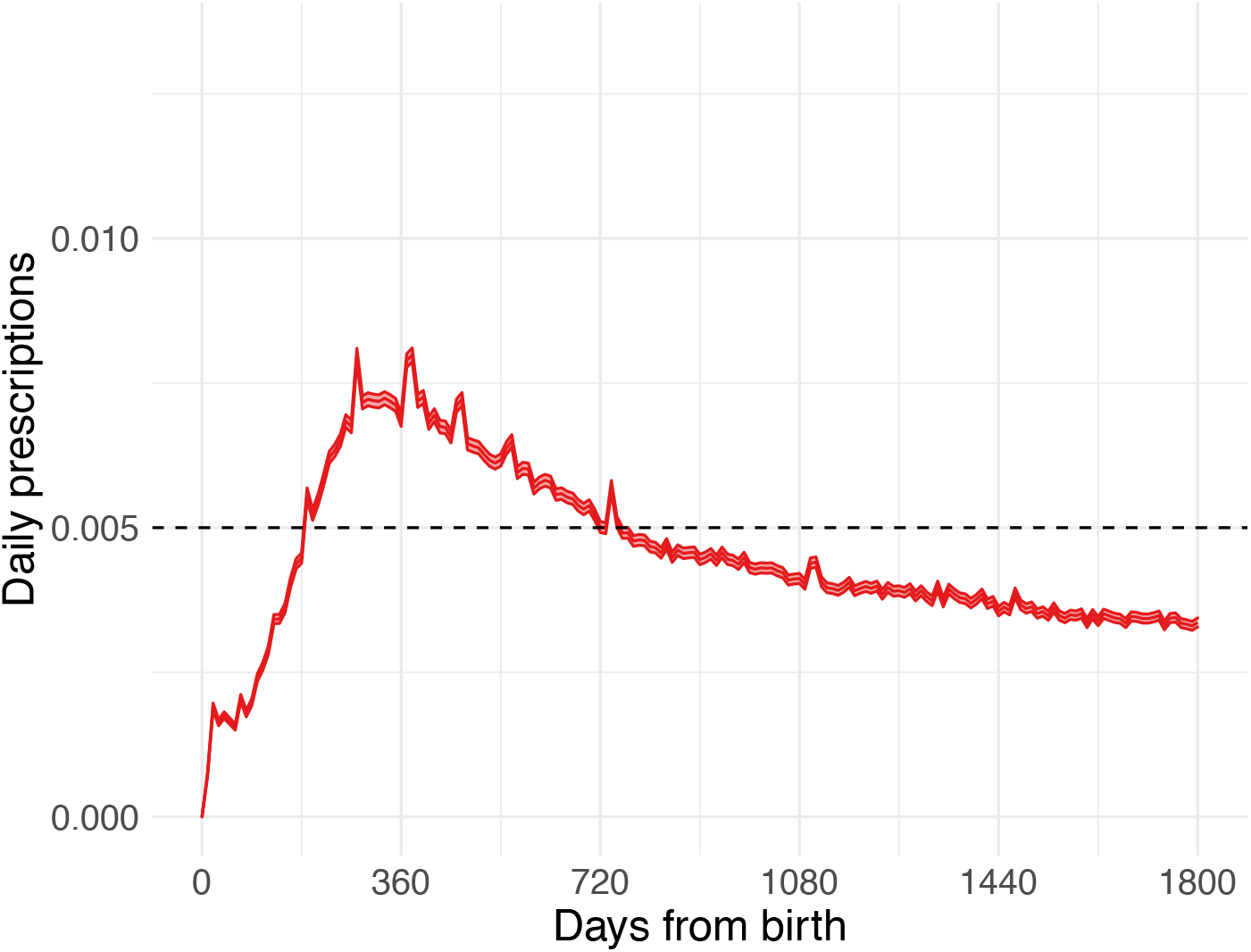
Mean daily antibiotic prescribing rates between birth and age 5. Daily rates are calculated by dividing the 30-day moving average of claims per person by 30. The solid line depicts the mean, and the shaded bands depict 95% confidence intervals for the mean according to a 2-tailed *t*-test for the mean. The dashed horizontal line at 0.005 marks the reference for 0.005 prescriptions per person per day or 0.15 prescriptions per person per 30 days.

**Supplementary Figure 2.**
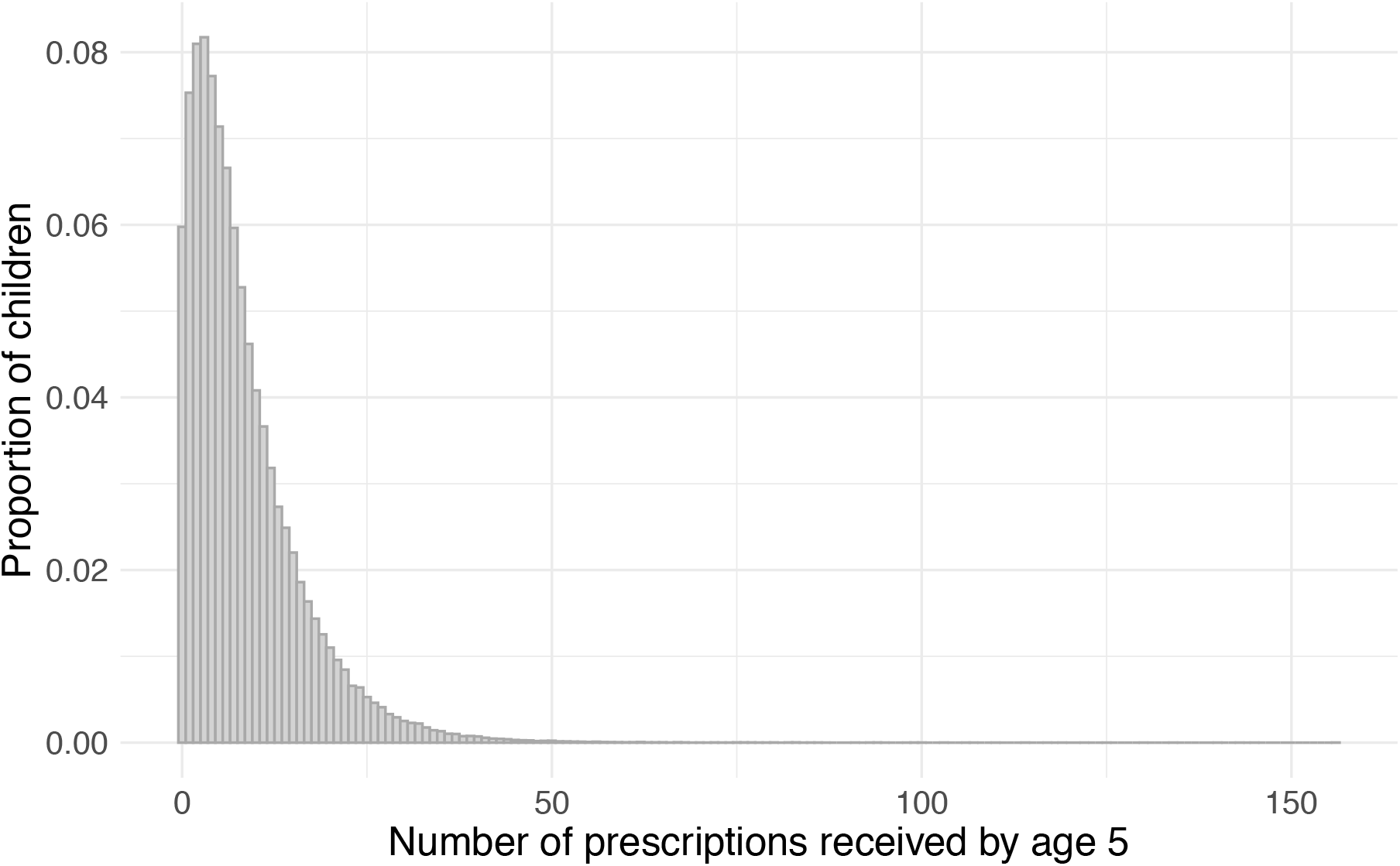
Histogram of the number of antibiotic prescriptions received by age 5 by children in the study population.

## References

1. Fleming-Dutra KE, Hersh AL, Shapiro DJ, Bartoces M, Enns EA, File TM, et al. Prevalence of inappropriate antibiotic prescriptions among us ambulatory care visits, 2010-2011. JAMA - J Am Med Assoc. 2016;315(17):1864–1873. doi:10.1001/jama.2016.4151

2. Lee GC, Reveles KR, Attridge RT, Lawson KA, Mansi IA, Lewis JS, et al. Outpatient antibiotic prescribing in the United States: 2000 to 2010. BMC Med. 2014;12(96). doi:10.1186/1741-7015-12-96

3. Korpela K, Salonen A, Virta LJ, Kekkonen RA, Forslund K, Bork P, et al. Intestinal microbiome is related to lifetime antibiotic use in Finnish pre-school children. Nat Commun. 2016;7(1):10410. doi:10.1038/ncomms10410

4. Bailey LC, Forrest CB, Zhang P, Richards TM, Livshits A, DeRusso PA. Association of Antibiotics in Infancy With Early Childhood Obesity. JAMA Pediatr. 2014;168(11):1063. doi:10.1001/jamapediatrics.2014.1539

5. Hicks LA, Bartoces MG, Roberts RM, Suda KJ, Hunkler RJ, Taylor TH, et al. US outpatient antibiotic prescribing variation according to geography, patient population, and provider specialty in 2011. Clin Infect Dis. 2015;60(9):1308–1316. doi:10.1093/cid/civ076

6. Lee GC, Reveles KR, Attridge RT, Lawson KA, Mansi IA, Lewis JS, et al. Outpatient antibiotic prescribing in the United States: 2000 to 2010. BMC Med. 2014;12(1):96. doi:10.1186/1741-7015-12-96

7. Poole NM, Shapiro DJ, Fleming-Dutra KE, Hicks LA, Hersh AL, Kronman MP. Antibiotic prescribing for children in United States emergency departments: 2009–2014. Pediatrics. 2019;143(2):2009–2014. doi:10.1542/peds.2018-1056

8. Nash DR, Harman J, Wald ER, Kelleher KJ. Antibiotic prescribing by primary care physicians for children with upper respiratory tract infections. Arch Pediatr Adolesc Med. 2002;156(11):1114–1119. doi:10.1001/archpedi.156.11.1114

9. Olesen SW, Barnett ML, MacFadden DR, Brownstein JS, Hernández-Díaz S, Lipsitch M, et al. The distribution of antibiotic use and its association with antibiotic resistance. Elife. 2018;7. doi:10.7554/eLife.39435

10. Ginsburg AS, Klugman KP. Vaccination to reduce antimicrobial resistance. Lancet Glob Heal. 2017;5(12):e1176–e1177. doi:10.1016/S2214-109X(17)30364-9

11. Klugman KP, Black S. Impact of existing vaccines in reducing antibiotic resistance: Primary and secondary effects. Proc Natl Acad Sci. 2018;115(51):12896–12901. doi:10.1073/pnas.1721095115

12. IBM Watson Health. IBM MarketScan Research Databases for Life Sciences Researchers.; 2019. https://www.ibm.com/us-en/marketplace/marketscan-research-databases

13. US Census Bureau. Metropolitan and Micropolitan. Published 2021. Accessed October 13, 2021. https://www.census.gov/programs-surveys/metro-micro.html

14. Stoecker C, Hampton LM, Moore MR. 7-Valent pneumococcal conjugate vaccine and otitis media: Effectiveness of a 2-dose versus 3-dose primary series. Vaccine. 2012;30(44):6256–6262. doi:10.1016/j.vaccine.2012.08.011

15. Agency for Healthcare Research and Quality. Clinical Classifications Software (CCS). Published 2019. Accessed September 9, 2021. https://www.hcup-us.ahrq.gov/toolssoftware/ccs10/ccs10.jsp

16. US Census Bureau. Explore Census Data. Published 2021. Accessed October 13, 2021. https://data.census.gov/cedsci/

17. Hobbs MR, Grant CC, Ritchie SR, Chelimo C, Morton SMB, Berry S, et al. Antibiotic consumption by New Zealand children: exposure is near universal by the age of 5 years. J Antimicrob Chemother. 2017;72(6):1832–1840. doi:10.1093/jac/dkx060

18. Fink G, D’Acremont V, Leslie HH, Cohen J. Antibiotic exposure among children younger than 5 years in low-income and middle-income countries: a cross-sectional study of nationally representative facility-based and household-based surveys. Lancet Infect Dis. 2020;20(2):179–187. doi:10.1016/S1473-3099(19)30572-9

19. Blue Cross Blue Shield. Antibiotic prescription fill rates declining in the U.S. Published online 2017. https://www.bcbs.com/sites/default/files/file-attachments/health-of-america-report/HoA.Antibiotics.Report.pdf

20. U.S. Department of Health and Human Services. Regional Offices. Published 2017. https://www.hhs.gov/about/agencies/iea/regional-offices/index.html

